# Associations of Trajectories of Loneliness and Neighborhood Stability with Depression, Alcohol and Substance Use, and Quality of Life among Women Living with HIV

**DOI:** 10.64898/2026.07.14.26358061

**Authors:** Peter Barr, Andrew Edmonds, Bradley Aouizerat, Mardge Cohen, Judith A. Cook, M. Reuel Friedman, Sabina Haberlen, Susan Holman, Mirjam-Colette Kempf, Deborah Konkle-Parker, Jennafer L. Kwait, David B. Hanna, Gayathri Pandey, Michael Plankey, Leah H. Rubin, Anna A. Rubtsova, Rebecca M. Schwartz, Azure B. Thompson, Deborah L. Jones, Jacquelyn L. Meyers, Tracey Wilson

**Author notes:** Corresponding author: Peter B. Barr,450 Clarkson Ave, MSC 1203, Brooklyn, NY 11203. These authors jointly supervised this work (Jacquelyn L. Meyers, Tracey Wilson).

## Abstract

Social relationships are an important social determinant of health. Loneliness, the perceived gap between one’s actual and desired relationships, has emerged as an important mechanism through which social relationships impact health. Like other intrapersonal-level factors associated with health, loneliness is influenced by broader social and structural factors, including characteristics of one’s neighborhood social environment. Although neighborhood-level protective and risk factors for loneliness and for mental health have been identified, prior studies have often focused solely on self-reported perceptions of the neighborhood environment. Further, few have considered aspects of the neighborhood social environments, such as neighborhood stability (i.e., stability of the community with long or short-term residents), independent of neighborhood socioeconomic conditions. In the current analysis, we explored longitudinal patterns of loneliness in conjunction with neighborhood stability among women with HIV (WWH) enrolled into the MACS/WIHS Combined Cohort Study (MWCCS) from 2014–2019 (*N*_2019_=1,394) to examine whether trajectories of loneliness and neighborhood stability were associated with depressive symptoms, non-prescription substance use, past-year cannabis use, number of alcoholic drinks per week, and several domains of quality of life. Loneliness at baseline (Betas = 0.24 – 0.54) and changes in loneliness over time (Betas = 0.11 – 0.26) were associated with each outcome, except for the association between changes in loneliness over time and drinks per week (Beta=0.13, *p* = 4.14×10^-2^), which did not persist after correcting for multiple comparisons. Neighborhood stability at baseline was associated with past year cannabis use (Beta=0.26, *p* = 1.00×10^-2^), depressive symptoms (Beta=-0.12, *p* = 1.54×10^-3^), and overall self-reported health (Beta=-0.08, *p* = 2.05×10^-2^). Changes in neighborhood stability across time were not associated with any outcome. Neighborhood stability moderated the association between changes in loneliness and general health perceptions. Our results demonstrate both overall loneliness and changes in loneliness over time have implications for current mental health in WWH, while changes in neighborhood stability did not.

## INTRODUCTION

Loneliness, defined as the negative emotional response that arises in response to perceptions that the number or quality of one’s social relationships does not meet needs or expectations, is an important determinant of health and well-being (Hawkley & Cacioppo, 2010; Holt-Lunstad, 2024; Office of the Surgeon, 2023; Power, Dolezal, Kee, & Lawlor, 2018; Wang et al., 2023). In the United States, over a third of adults report feeling lonely sometimes or always, with higher prevalence among individuals living with chronic health conditions and those of lower socioeconomic status (Albertorio-Diaz & Wheldon, 2025).

Longitudinal studies show that greater loneliness is associated with a variety of adverse outcomes, including lower perceived coping ability, increased psychological distress, and among older adults, increased risk of cognitive decline and frailty (Brouillette et al., 2022; Han et al., 2017; Meireles et al., 2023; Rubtsova et al., 2021). Importantly, people with HIV (PWH) tend to report higher levels of loneliness as compared to the general population (Mazonson et al., 2021; Pollak, Cotton, Winter, & Blumen, 2025). However, research on loneliness among PWH has been predominantly cross-sectional and disproportionately focused on men (Pollak et al., 2025), despite emerging evidence of gender differences in loneliness-related outcomes. For instance, in response to COVID-19 related social disruptions, women with HIV (WWH) were more likely than men to report increased loneliness (Jones et al., 2021), and some research suggests that among PWH, the association between loneliness and alcohol or substance use may be stronger for women as compared to men (Mannes et al., 2016). Understanding how the burden of loneliness over time is associated with behavioral health outcomes and with HIV-related quality of life among women is important, given that chronic loneliness may be a more important determinant of health outcomes than transient or temporary feelings of loneliness (Hajek et al., 2025).

Loneliness is shaped not only by individual factors and interpersonal dynamics, but by broader social and structural conditions, including characteristics of the neighborhoods in which people live (Feng & Astell-Burt, 2022). Access to green space, walkability, availability of community resources, and lower neighborhood-level socioeconomic deprivation have all been linked to less loneliness (Schepers et al., 2025), and many of these same neighborhood characteristics have been associated with engagement in HIV care (Kerr et al., 2024; Kimaru et al., 2024).

Much of the research on neighborhood factors and associations with loneliness and health utilize a resource-availability perspective to understand how adverse neighborhood environments influence health outcomes. They examine relationships between neighborhood characteristics and health at a single point in time. However, even when changes in neighborhoods bring about improvements in safety, services, or other attributes that reduce area-level social inequalities, such changes can reduce social cohesion and contribute to loneliness (Arcaya et al., 2024). The construct of neighborhood stability reflects how frequently residents are replaced within a defined area and time period, measured through area-level indices. Typically, they are measured using the percentage of people in a census tract who have resided in the same location for the past five years (Ross et al., 2000; Schieman, 2009). Neighborhood stability is a measure of residential consistency and mobility, related but distinct from area-level socioeconomic status, and can occur in situations of increasing neighborhood disadvantage as well as area-level growth and gentrification.

The degree to which neighborhood stability influences the health of residents may vary depending on individual characteristics and does not affect everyone equally (Haslam, Fong, Haslam, & Cruwys, 2024). For some populations, including older adults and those with chronic illness, disability, or poor health, the stability of one’s neighborhood may be particularly tied to health and well-being (Goldman et al., 2023). The press-competence model, for instance, describes how human functioning is determined by both an individual’s perceived intrapersonal physical, social, and psychological competencies and the physical, social, and psychological demands present in a given geographic area, described as environmental press. Older people and those with illness-associated limitations in physical or cognitive function may have reduced capacity to respond to changes in neighborhood environments, including changes in social networks, in turn resulting in increased stress and reductions in well-being and quality of life (Lam & Baxter, 2025; Park et al., 2019; Scheidt et al., 2003; Wahl & Lang, 2003).

To date, little research has examined how neighborhood stability contributes to loneliness among WWH or whether neighborhood stability and loneliness independently or jointly shape HIV-related quality of life or known behavioral health indices, such as alcohol and substance use and mental health, that are known to contribute to quality of life among WWH (Waldron et al., 2021; Yang et al., 2018). We worked with participants of the Women’s Interagency HIV Study (WIHS) to examine the joint influence of neighborhood stability and loneliness on multiple mental health outcomes.

## METHODS

### Participants

The data analytic sample included participants in the Women’s Interagency HIV Study (WIHS) who were living with HIV in 2014, when the study began to implement measures of loneliness, and who were active in the study between 2014 and 2019. WIHS clinical study sites during this period included Bronx, NY; Brooklyn, NY; Washington, DC; San Francisco, CA; Chicago, IL; Atlanta, GA; Miami, FL; Chapel Hill, NC; Birmingham, AL/Jackson, MS. Participants were seen at 6-month intervals for physical assessments, specimen collection, and structured interviews; home addresses were collected annually. All study procedures were approved by institutional review boards at participating sites and written informed consent was obtained from all participants (Adimora et al., 2018). The WIHS merged with a similar men’s study, the Multicenter AIDS Cohort Study (MACS) in 2019, and is now known as the MACS/WIHS Combined Cohort Study (MWCCS).

The current analyses utilize data derived from annual visits in which participants were administered questions on loneliness (2014-2019; 6 time points in total). Our final analytic sample was limited to WWH who responded during the 2019 assessment, regardless of the number of observations they provided between 2014 and 2019 (N=1,392; mean number of observations per person=5.7).

### Measures

Loneliness was measured with the 3-item version of the UCLA Loneliness Scale (Hughes et al., 2004; Russell et al., 1978). For each annual time point, the value for loneliness was taken as the mean of the three composite items: 1) “How often do you feel that you lack companionship?”, 2) “How often do you feel left out?”, and 3) “How often do you feel isolated from others?”. Responses ranged from 1 (“Hardly ever”) to 3 (“Often”). We scaled items from 0 to 1 for model convergence in the longitudinal analyses.

To assess neighborhood stability, each participant’s home address was first geocoded to its corresponding census tract, and then to 5-year estimates of indicators from the American Community Survey (ACS). We chose 5-year estimates for each time point as these are more reliable than the 1-year estimates. Neighborhood stability is typically measured using the percentage of people in a tract who have resided in the same location for the past five years (Ross et al., 2000; Schieman, 2009). However, the ACS asks about the percentage of people in a tract who have resided in the same location for varying time frames across years. Therefore, to measure neighborhood stability over time, we selected three correlated ACS measures defined as relevant for neighborhood stability in previous research (Cohen & Pettit, 2019): % of homes that are owner occupied, % of homes that are vacant, and % of rental units that are vacant. We calculated an index of neighborhood stability by summing items and converting to Z-scores, with greater values indicating greater neighborhood stability at yearly intervals from 2014 to 2019.

We considered twelve outcomes, all of which were abstracted at the 2019 visit. Outcomes included: 1) depressive symptoms using the Center for Epidemiologic Studies Depression scale (CES-D) (Radloff, 1977) (range 0–60, with higher scores indicating more severe depressive symptoms); 2) number of alcoholic drinks per week, 3) any cannabis use since last visit, 4) use of any other substances, including cocaine, crack, heroin, methamphetamine, or non-prescribed methadone since last visit, and 8 HIV-related quality of life (QOL) domains assessed using the AIDS Clinical Trials Group (ACTG) SF-21 (Wu et al., 1997), including 1) emotional well-being, 2) energy and fatigue, 3) cognitive function, 4) bodily pain, 5) role functioning, 6) social functioning, 7) physical functioning, and 8) general health perception; each domain was scored from 0 (lowest QOL) to 100 (highest QOL). ACTG SF-21 was originally adapted from the Medical Outcomes Study HIV Health Survey (MOS-HIV) and has established reliability and validity in populations with HIV (Wu et al., 1997). Variables were reverse-coded if necessary, so that higher scores indicated worse functioning or poorer outcomes.

Because neighborhood stability is associated with changes in neighborhood socioeconomic conditions, we created two covariates from the 2014 5-year ACS estimates: 1) an index of neighborhood disadvantage (defined as the percentage of the total population at or below the federal poverty threshold, percentage of adults that are unemployed, percentage of female-headed households with children); and 2) an index of neighborhood advantage (defined as the percentage of adults over 25 with a college degree, median household income, and percentage of adults with a professional occupation) in line with prior neighborhood analyses (Barr, 2018a, 2018b). Each of the indices was converted to Z-scores for interpretation and included as a covariate. Additional demographic covariates relevant for mental health included age (at 2019), race and ethnicity, relationship status (i.e., widowed or divorced, married or living with a partner, other relationship status), and education level (i.e., did not complete high school, completed high school or higher).

### Analytic Strategy

First, we created baseline values and indicators for change over time for loneliness and neighborhood stability using Empirical Bayes (EB) estimates from linear mixed effects models. The mixed model is useful in that it is flexible, allowing us to include participants regardless of 1) the number of responses they provided or 2) their schedule of responses across the course of the study (Singer & Willett, 2003). For each model, we first inspected mean values to visually explore the functional form of change over time. Next, we fit a series of nested models to identify the most appropriate model in terms of random effects and fixed effects estimates. After identifying the best fitting model, we calculated the posterior means of the random effect and carried those values forward into the association analyses with our outcomes of interest. In the main analyses, we first examined the bivariate association between each measure included in the longitudinal models and each outcome using linear or logistic models for continuous and binary outcomes, respectively. Next, we estimated a fully adjusted model, including all predictors simultaneously, as well as including age, race and ethnicity, relationship status, education, index of neighborhood advantage, and index of neighborhood disadvantage as covariates. To correct for multiple testing, we applied a false discovery rate (FDR) of 5% to all analyses (Benjamini & Hochberg, 1995).

## RESULTS

Study participants had a mean age of 52.5yrs (SD=9.2) as of the visit in 2019, with the majority of respondents identifying as Black or African American (62.3%) and having completed high school or equivalent (57.1%). In terms of outcomes, respondents reported drinking 2.1 drinks per week (SD=6.4); on average, 21.1% respondents indicated using cannabis since the last visit, and 5.9% reported some other form of substance use (Table 1). Participants reported a mean CES-D score of 11.6 (SD=11.2), with 412 respondents (29.7%) reporting scores indicative of clinical depression (e.g., ≥ 16) (Lewinsohn et al., 1997). For HIV-related QOL domains, mean scores were 68.4 (SD=28.1) for bodily pain, 72.2 (SD=23.5) for emotional well-being, 59.9 (SD=26.1) for energy, 78.2 (SD=28.2) for role functioning, 78.6 (SD=26.1) for social functioning, 68.8 (SD=29.5) for physical functioning, 84.0 (SD=23.4) for cognitive functioning, and 68.3 (SD=19.8) for perception of overall health. Almost all women were on HIV antiretroviral therapy (91.7%); 74.1% had an HIV RNA viral load <200 copies/mL, and 71.9% CD4+ cells/mL^3^ of 500 or greater.

**Table 1:** Characteristics of WWH (N = 1,392)

|  | <u>Mean/N*</u> | <u>SD/%*</u> | <u>Min</u> | <u>Max</u> |
| --- | --- | --- | --- | --- |
| Number of neighborhood observations | 5.69 | 0.64 | 1 | 6 |
| Age (at 2019 visit), years | 52.46 | 9.15 | 29 | 84 |
| Race and ethnicity: |  |  |  |  |
| Non-Hispanic Black/African American | 839 | 60.28% | - | - |
| Non-Hispanic White | 131 | 9.48% | - | - |
| Hispanic or Latino/a/x | 103 | 7.40% | - | - |
| Other or multi racial-ethnic identity | 315 | 22.63% | - | - |
| Education: |  |  |  |  |
| Less than high school | 427 | 30.68% | - | - |
| High school or some college | 795 | 57.11% | - | - |
| College graduate or more | 112 | 8.05% | - | - |
| Relationship status: |  |  |  |  |
| Married or living with a partner | 355 | 25.50% | - | - |
| Never married | 449 | 32.26% | - | - |
| Divorced, separated, or widowed | 400 | 28.74% | - | - |
| Other | 115 | 8.26% | - | - |
| Outcomes: |  |  |  |  |
| Cannabis use (since last visit) | 294 | 21.12% | - | - |
| Other drug use (since last visit) | 82 | 5.89% | - | - |
| Drinks per week | 2.08 | 6.39 | 0 | 77 |
| Depressive symptoms (CES-D) | 11.59 | 11.21 | 0 | 54 |
| Emotional well-being | 72.19 | 23.5 | 0 | 100 |
| Energy/Fatigue | 59.89 | 26.06 | 0 | 100 |
| Role functioning | 78.21 | 28.15 | 0 | 100 |
| Social functioning | 78.58 | 26.06 | 0 | 100 |
| Physical functioning | 68.81 | 29.54 | 0 | 100 |
| Cognitive functioning | 83.97 | 23.41 | 0 | 100 |
| General health perception | 68.29 | 19.84 | 0 | 100 |
| Bodily pain | 68.35 | 28.14 | 0 | 100 |

\* mean/N and SD% based on all participants without missing data

Next, we fit a series of growth models for the trajectories of loneliness and neighborhood stability using linear mixed models. Figure 1 presents the mean estimates for loneliness (N=8,484 total person-years) and neighborhood stability (N=7,464 total person-years) over the span of the yearly visits from 2014 to 2019. Visual inspection suggested that a linear function fit the patterns in change over time for both loneliness and neighborhood stability. Table 2 presents the results from the nested model comparisons for each measure. Overall, the model fitting results confirmed that a linear change was the best fit for both. For loneliness, the intraclass correlation coefficient (ICC) was relatively strong (0.54), suggesting that the majority of the variance in loneliness over time was due to within-person factors. Compared to the unconditional models (intercept-only), the linear growth model fit significantly better (Δχ*^2^*=4.38, df=1, *p*=3.63 x 10^-2^), and a quadratic function did not improve model fit (Δχ*^2^* 0.48, df=1, *p*=4.89 x 10^-1^). The inclusion of a random slope for time notably improved model fit (Δχ*^2^*=67.15, df=2, *p*=2.62 x 10^-15^). Neighborhood stability had a similar pattern of results, with a strong ICC (0.81), the linear growth model fitting significantly better (Δχ*^2^*=80.17, df=1, *p*=3.43 x 10^-19^), a quadratic function not improving model fit (Δχ*^2^*=2.26, df=1, *p*=1.32 x 10^-1^), and the addition of the random slope for time yielding the best fit overall (Δχ*^2^*=971.32, df=2, *p*=1.20×10^-211^). From these final models that included a linear effect for time, a random effect for the slope of time, and random intercepts, we generated individual estimates for their “intercept” (deviation from the average value at 2014 visit) and “slope” (deviation from the average rate of change across visits from 2014 through 2019) in both loneliness and neighborhood stability.

**Figure 1:**
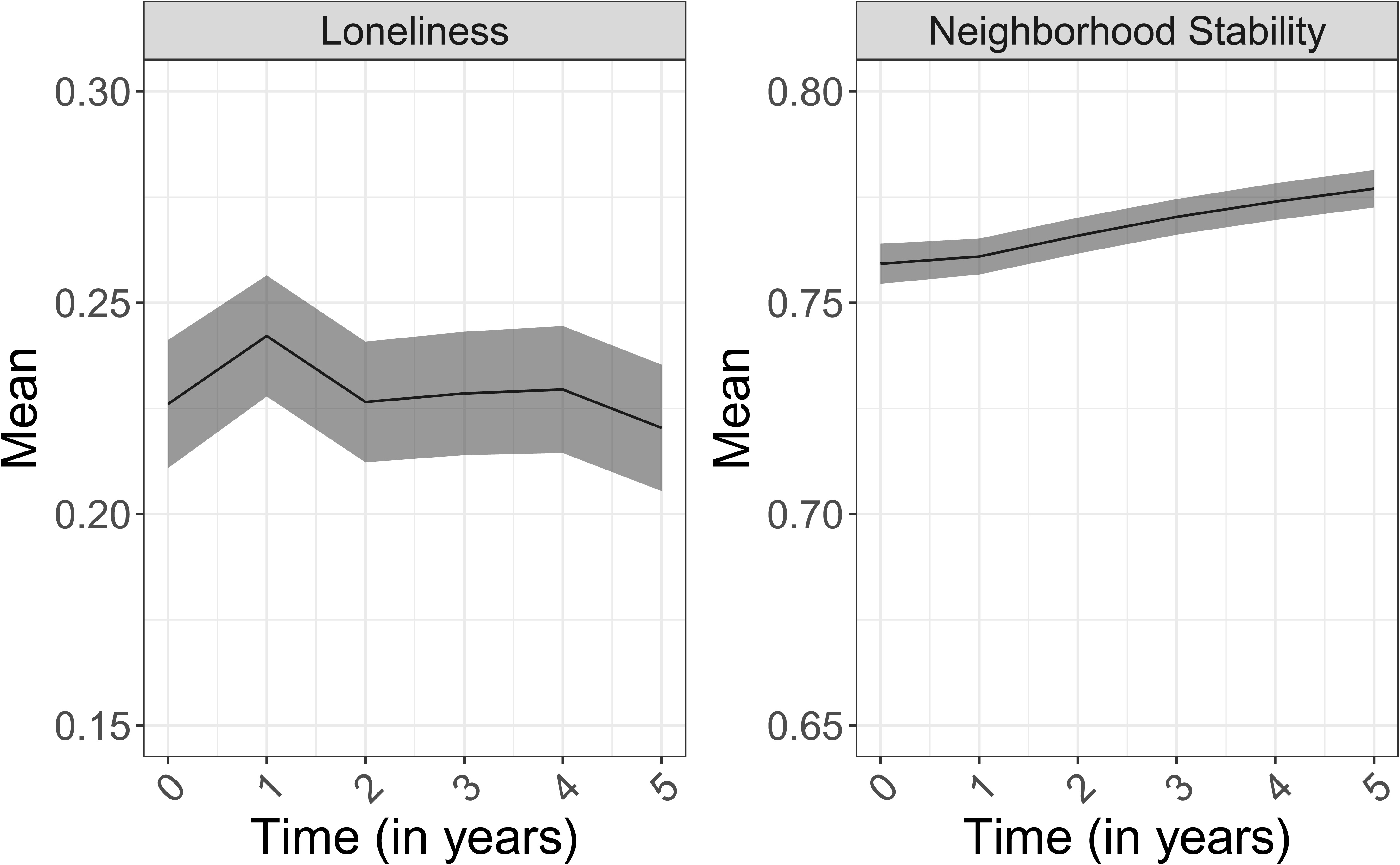
*Mean loneliness and neighborhood stability over time (2014-2019).* Mean levels of loneliness and neighborhood stability (and corresponding 95% confidence intervals) from annual assessments across 2014 to 2019.

**Table 2:** Model fit for longitudinal growth models.

| <u>Outcome</u> | <u>Model</u> | <u>N par</u> | <u>Comparison</u> | <u>AIC</u> | <u>BIC</u> | <u>LL</u> | <u>Δχ<sup>2</sup></u> | <u>Δdf</u> | <u>P-value</u> |
| --- | --- | --- | --- | --- | --- | --- | --- | --- | --- |
| Loneliness | 1. Unconditional growth model | 3 | - | 46.43 | 67.57 | -20.22 | - | - | - |
| Loneliness | 2. Linear growth model | 4 | Model 1 | 44.05 | 72.23 | -18.02 | 4.38 | 1 | 3.63 x 10<sup>-2</sup> |
| Loneliness | 3. Quadratic growth model | 5 | Model 2 | 45.57 | 80.80 | -17.79 | 0.48 | 1 | 4.89 x10<sup>-1</sup> |
| Loneliness | 4. Linear growth model + random slope | 6 | Model 2 | -19.10 | 23.18 | 15.55 | 67.15 | 2 | 2.62 x10<sup>-15</sup> |
| Neighborhood stability | 1. Unconditional growth model | 3 | - | -22851.56 | -22830.80 | 11428.78 | - | - | - |
| Neighborhood stability | 2. Linear growth model | 4 | Model 1 | -22929.73 | -22902.06 | 11468.87 | 80.17 | 1 | 3.43 x10<sup>-19</sup> |
| Neighborhood stability | 3. Quadratic growth model | 5 | Model 2 | -22930.00 | -22895.41 | 11470.00 | 2.26 | 1 | 1.32 x10<sup>-1</sup> |
| Neighborhood stability | 4. Linear growth model + random slope | 6 | Model 2 | -23897.05 | -23855.54 | 11954.53 | 971.32 | 2 | 1.20 x10<sup>-211</sup> |

N par = number of parameters in the model; AIC = Akaike's information criteria; BIC = Bayesian information criteria; LL = log-likelihood; df = degrees of freedom

Next, we examined associations between the intercepts and slopes for loneliness and neighborhood stability derived from the longitudinal models for each outcome. Figure 2 presents fully adjusted results (conditional estimates adjusted for other predictors and sociodemographic covariates, bivariate results presented in supplementary material) for associations with each outcome. All predictors and (continuous) outcomes were transformed to Z-scores, so parameters can be interpreted as a one standard deviation change in predictor corresponding to a one standard deviation change in outcome. Loneliness at the beginning of the observation period (intercept) was positively associated (hereafter shown as Beta/Odds Ratio [95% CI]) with alcohol use (Beta=0.279 [0.142, 0.417]), cannabis use (OR=1.427 [1.242, 1.639]), other substance use (OR=1.594 [1.284, 1.979]), and depressive symptoms (Beta=0.543 [0.488, 0.599]). For QOL domains, baseline loneliness was positively associated with pain (Beta=0.334 [0.281, 0.388]), poor emotional well-being (Beta=0.512 [0.463, 0.562]), and fatigue (Beta=0.332 [0.278, 0.386]). Baseline loneliness was also positively associated with poor role functioning (Beta=0.281 [0.226, 0.336]), social functioning (Beta=0.428 [0.377, 0.478]), physical functioning (Beta=0.243 [0.188, 0.297]), and cognitive (Beta=0.402 [0.350, 0.454]) functioning. Finally, baseline loneliness was positively associated with poor perceptions of general health (Beta=0.405 [0.353, 0.457]).

**Figure 2:**
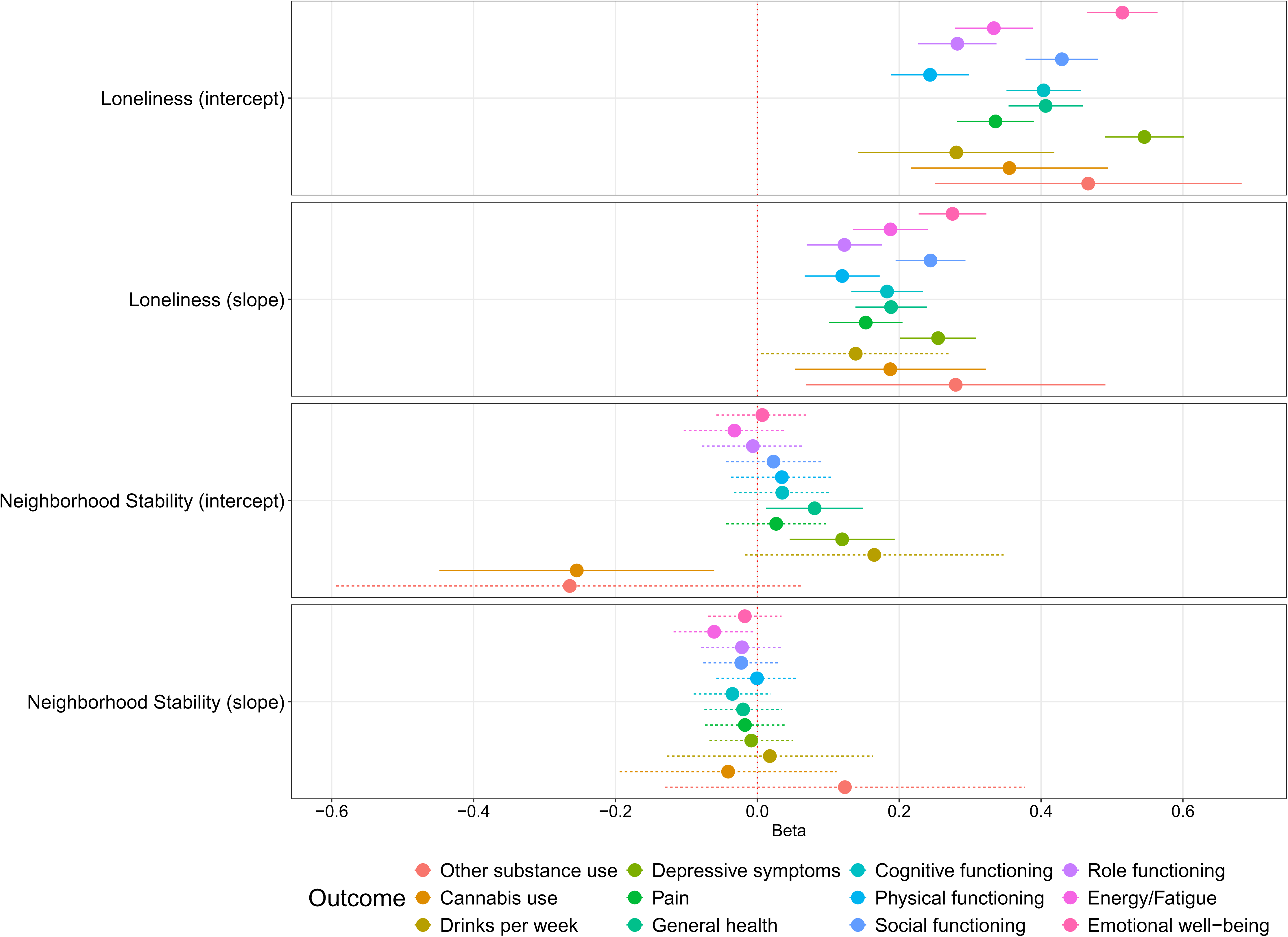
*Associations between loneliness, neighborhood stability, and mental health outcomes.* Adjusted betas and log(odds ratios) and corresponding 95% confidence intervals from linear and logistic models, respectively. Solid lines represent significant associations after correcting for multiple testing. All models included age (at 2019), race-ethnicity, relationship status, education, neighborhood advantage, and neighborhood disadvantage as covariates.

There was a similar pattern of results for changes in loneliness over time (slope) in the fully adjusted models. Increases in loneliness over the course of the observation period were positively associated with cannabis use (OR=1.206, [1.054, 1.380]), other substance use (OR=1.323, [1.071, 1.634]), depressive symptoms (0.239, [0.189, 0.289]), pain (0.143, [0.095, 0.192), poor emotional well-being (Beta=0.258, [0.213, 0.302]), fatigue (Beta=0.176, [0.127, 0.225]), poor role functioning (Beta=0.115, [0.065, 0.165]), and poor social functioning (Beta=0.229, [0.183, 0.275]), poor physical functioning (Beta=0.112, [0.063, 0.162]), and poor cognitive functioning (Beta=0.171, [0.124, 0.219]). Increased loneliness was also positively related to poorer perceptions of general health (Beta=0.177, [0.130, 0.224]). Only alcoholic drinks per week (since last visit) was unrelated to the slope of loneliness across the observation period. Importantly, while the pattern of results was similar to that for loneliness at baseline, the effect sizes for change in loneliness were markedly smaller. Lastly, it is worth noting that there were minimal changes in effect sizes between the bivariate results to those from the multivariable analyses, suggesting that these associations were minimally affected by differences in sociodemographic or neighborhood socioeconomic characteristics.

Neighborhood stability at the beginning of the observation period was negatively associated with cannabis use since last visit (OR=0.775, [0.639, 0.941]) but positively associated with depressive symptoms (Beta=0.120, [0.046, 0.194]) and poor perceptions of overall health (Beta=0.081, [0.013, 0.149]). None of the other outcomes were associated with neighborhood stability at baseline. Additionally, there were no significant associations with changes in neighborhood stability.

Lastly, we tested for interactions between each of the loneliness and neighborhood stability predictors, as well as for their interactions with age. We found no evidence for any interactions with age (full results in supplemental material). However, there were two interactions that survived correction for multiple testing for general health perceptions presented, in Figure 3, 1) change in loneliness and change in neighborhood stability (Beta=0.084, p=1.76×10^-3^), and 2) change in loneliness and baseline neighborhood stability (Beta=0.085, p=2.99 x10^-3^). Experiences of increase in loneliness over time were particularly relevant for those from high stability neighborhoods at baseline, but not as neighborhood stability decreased. Similarly, as neighborhood stability increased over time, the association between changes in loneliness and poor health perceptions became stronger at the 2019 visit.

**Figure 3:**
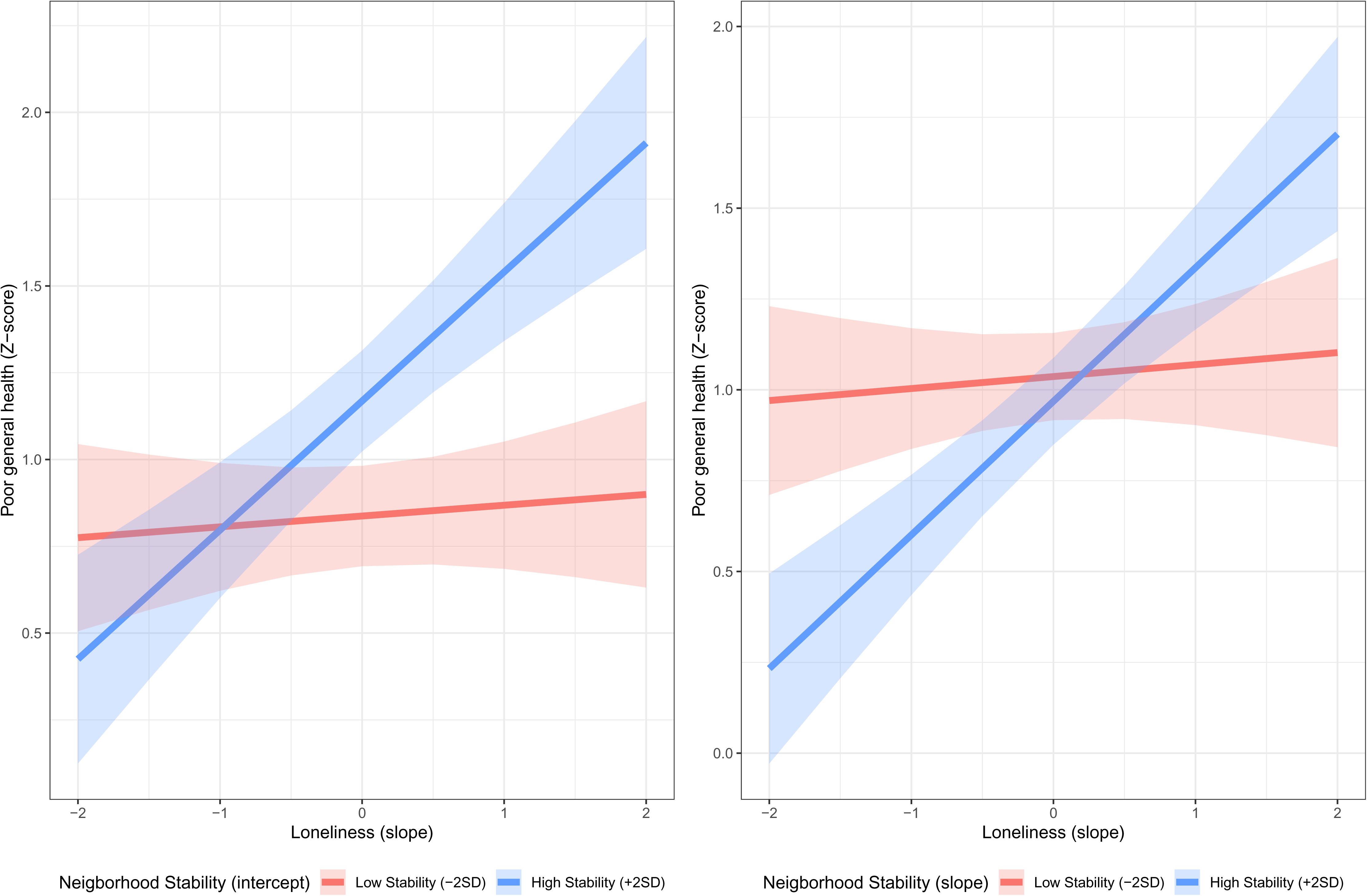
*Poor General Health across levels of Neighborhood Stability and Changes in Loneliness.* Predicted values (and 95% confidence intervals) of poor physical functioning for interactions between: 1) neighborhood stability (intercept) and loneliness (slope) on the left, and 2) neighborhood stability (slope) and loneliness (slope) on the right. All values presented as Z-scores for interpretation.

## DISCUSSION

Social relationships are important determinants of health and well-being. Loneliness is a key mechanism linking social relationships and health. In people living with HIV, social isolation and social support seem to be especially important, as they are associated with multiple aspects of mental health, substance use, and psychological functioning (Friedman et al., 2017; Greene et al., 2018; Pollak et al., 2025), and these can have profound impacts on disease management. Like all proximal social determinants of health, social relationships are also patterned by other social factors, including the neighborhood social environment. In the current analysis, we examined the degree to which longitudinal changes in loneliness and neighborhood stability were associated with severity of depressive symptoms, alcohol and substance use, and multiple quality of life measures in WWH.

Prior loneliness and its change over time had profound consequences for current reports of mental and emotional well-being in WWH. Importantly, results from fully adjusted models revealed that loneliness five years ago as well as the trajectory of loneliness over the preceding five years led to poorer health and behavioral consequences. These results align with prior work on loneliness and other domains of social relationships. For example, research on social interactions using a longitudinal, ecological momentary assessment approach found that more daily social interactions were associated with higher ratings of real-time happiness (Kamalyan et al., 2022). Using data from the Crisis Text Line from 2016-2022, researchers identified co-occurrence between social isolation and loneliness with depression/sadness, anxiety, and suicidality (Lucero et al., 2024). These findings are consistent with those from the current analysis, demonstrating the importance of exploring loneliness in a longitudinal framework. Lastly, it is worth noting that there were minimal changes in effect sizes between bivariate and fully adjusted results, suggesting that these associations were minimally affected by differences in sociodemographic or neighborhood socioeconomic characteristics.

Our measure of neighborhood stability was developed to reflect the ability of residents to form or maintain relationships, potentially contributing to loneliness. While neighborhood stability at baseline was negatively associated with prior-year cannabis use, it was not associated with reports of other substance use or level of alcohol use, nor was it associated with emotional well-being, pain, energy/fatigue, role functioning, social functioning, cognitive health, or mental health. We did not detect associations with longitudinal changes in neighborhood stability. Perhaps counterintuitively, the only other significant associations detected were negative relationships between greater neighborhood stability at baseline with higher burden of depressive symptoms and lower general health perceptions.

Prior work has shown that short-term (but not long-term) increases in neighborhood vacancy were associated with poorer mental health, after adjustment for individual covariates (Pearson et al., 2019). This could explain our finding of associations between neighborhood vacancy and poor outcomes at baseline but not over the long term. It could also be that declining residential occupancy is only indirectly related to negative outcomes and that other factors such as social integration and sense of community are more directly tied to outcomes, as also found in the Flint study. Perhaps counterintuitively, the only other significant associations were negative relationships between neighborhood stability at baseline and current depressive symptoms and general health perceptions (reverse coded). Prior research has consistently found associations between neighborhood social conditions and psychological distress in the opposite direction (Barr, 2018a; Hill et al., 2005; Ross, 2000; Ross & Mirowsky, 2001, 2009).

Though we did not find much evidence for main effects of neighborhood stability, we did find evidence of interactions between changes in loneliness and neighborhood stability (both baseline and change over time) for general health perceptions, such that changes in loneliness were particularly relevant for those from neighborhoods that were: 1) highly stable at baseline, and 2) increased in stability over time. These results help demonstrate the potentially contextual nature of loneliness and that for those in highly unstable neighborhoods, other factors may be more relevant, at least regarding general health perceptions.

The null results for neighborhood stability could also reflect the fact that our operationalization of stability captures residential occupancy rather than neighborhood social environment. Neighborhood conditions related to inclusivity, trust, belonging, and shared spaces may be more important, or may interact with other social environmental conditions. For example, in a study of post-industrial neighborhood decline in Flint, Michigan, increases in vacancy were associated with poorer mental health outcomes after controlling for individual-level demographic characteristics, but this effect was only detected among people with lower social ties to their neighbors (Pearson et al., 2019). Future analyses in samples that include both census-derived measures and perceptions of neighborhood social environment will allow us to determine which domains are more relevant for health and well-being.

Research on changes in indicators of neighborhood conditions have been primarily limited to studies of perceptions of change or stability, rather than utilizing neighborhood-level indicators of neighborhood stability. Persons with lower quality of life, greater loneliness, or common mental health conditions may be more likely to perceive their environment as having negative characteristics (Lyu & Forsyth, 2021). Clarifying these relationships utilizing geospatial measures of neighborhood stability is therefore critical to creating a comprehensive picture of neighborhood shifts to inform policies to support social, emotional, and physical well-being. Further research examining both objective and subjective indices of neighborhood factors over time and their interactions may help further clarify their role in mental health and quality of life.

This work has several limitations. First, the sample is comprised entirely of WWH enrolled in a prospective cohort study and follows a population who are older than the national average of WWH and at a limited number of geographic clinical research sites (D’Souza et al., 2021). As such, it is not known whether these results would generalize to men living with HIV or to the broader population of WWH in the US. Second, while ACS data have the benefit of being unconfounded in contrast to observations from self-report, they may not capture all the relevant aspects of neighborhood social environment. Future work should consider methods of aggregating self-reported observations within neighborhoods to balance self-report with reporter bias (Mujahid et al., 2007; Raudenbush & Sampson, 1999).

Aspects of social relationships, especially loneliness, are key determinants of health. Our results demonstrate that above and beyond prior levels of loneliness, changes in loneliness over time are independently related to several aspects of quality of life and behavioral and mental health among WWH. Some approaches, such as psychological interventions and social and emotional skills training, have been shown to produce small to moderate effects on reducing loneliness (Lasgaard et al., 2022). However, research on their effectiveness among populations with HIV is lacking (Zhao et al., 2025). These findings support the importance of continued innovation in the design and implementation of effective interventions to prevent and reduce loneliness in this population.

## Data Availability

Data are available through controlled release. More information on data access can be found here: https://statepi.jhsph.edu/mwccs/data-sharing/

## ACKNOWLEDGEMENT

This work was supported through the National Institute of Mental Health, R01MH128955 (Tracey Wilson and Jacquelyn Meyers). Data in this manuscript were collected by the Women’s Interagency HIV Study (WIHS), now the MACS/WIHS Combined Cohort Study (MWCCS). The contents of this publication are solely the responsibility of the authors and do not represent the official views of the National Institutes of Health (NIH). MWCCS (Principal Investigators): Atlanta CRS (Cecile Lahiri, Anandi Sheth, and Gina Wingood), U01-HL146241; Bronx CRS (David Hanna and Anjali Sharma), U01-HL146204; Brooklyn CRS (Deborah Gustafson and Tracey Wilson), U01-HL146202; Data Analysis and Coordination Center (Gypsyamber D’Souza, Stephen Gange and Elizabeth Topper), U01-HL146193; Chicago-Cook County CRS (Mardge Cohen, Audrey French, and Ryan Ross), U01-HL146245; Northern California CRS (Bradley Aouizerat, Jennifer Price, and Phyllis Tien), U01-HL146242; Metropolitan Washington CRS (Seble Kassaye and Daniel Merenstein), U01-HL146205; Miami CRS (Maria Alcaide, Claudia Martinez, and Deborah Jones), U01-HL146203; UAB-MS CRS (Mirjam-Colette Kempf, James B. Brock, and Emily Levitan), U01-HL146192; UNC CRS (M. Bradley Drummond and Michelle Floris-Moore), U01-HL146194. The MWCCS is funded primarily by the National Heart, Lung, and Blood Institute (NHLBI), with additional co-funding from the Eunice Kennedy Shriver National Institute of Child Health & Human Development (NICHD), National Institute on Aging (NIA), National Institute of Dental & Craniofacial Research (NIDCR), National Institute of Allergy and Infectious Diseases (NIAID), National Institute of Neurological Disorders and Stroke (NINDS), National Institute of Mental Health (NIMH), National Institute on Drug Abuse (NIDA), National Institute of Nursing Research (NINR), National Cancer Institute (NCI), National Institute on Alcohol Abuse and Alcoholism (NIAAA), National Institute on Deafness and Other Communication Disorders (NIDCD), National Institute of Diabetes and Digestive and Kidney Diseases (NIDDK), National Institute on Minority Health and Health Disparities (NIMHD), and in coordination and alignment with the research priorities of the National Institutes of Health, Office of AIDS Research (OAR). MWCCS data collection is also supported by UL1-TR000004 (UCSF CTSA), UL1-TR003098 (JHU ICTR), UL1-TR001881 (UCLA CTSI), P30-AI-050409 (Atlanta CFAR), P30-AI-073961 (Miami CFAR), P30-AI-050410 (UNC CFAR), P30-AI-027767 (UAB CFAR), P30-AI-124414 (ERC-CFAR), P30-MH-116867 (Miami CHARM), UL1-TR001409 (DC CTSA), KL2-TR001432 (DC CTSA), and TL1-TR001431 (DC CTSA). The authors gratefully acknowledge the contributions of the study participants and dedication of the staff at the MWCCS sites and of the Social Connections Community Advisory Group (Jeanette Carter, Marta Santiago, Marc C.E. Wagner, Martha Williams).

## REFERENCES

Adimora, A. A., Ramirez, C., Benning, L., Greenblatt, R. M., Kempf, M.-C., Tien, P. C., Kassaye, S. G., Anastos, K., Cohen, M., Minkoff, H., Wingood, G., Ofotokun, I., Fischl, M. A., & Gange, S. (2018). Cohort Profile: The Women’s Interagency HIV Study (WIHS). International Journal of Epidemiology, 47(2), 393–394i. 10.1093/ije/dyy021

Arcaya, M. C., Ellen, I. G., & Steil, J. (2024). Neighborhoods And Health: Interventions At The Neighborhood Level Could Help Advance Health Equity. Health Affairs, 43(2), 156–163. 10.1377/hlthaff.2023.01037

Barr, P. B. (2018a). Early neighborhood conditions and trajectories of depressive symptoms across adolescence and into adulthood. Advances in Life Course Research, 35, 57–68. 10.1016/j.alcr.2018.01.005

Barr, P. B. (2018b). Neighborhood conditions and trajectories of alcohol use and misuse across the early life course. Health and Place, 51, 36–44. 10.1016/j.healthplace.2018.02.007

Benjamini, Y., & Hochberg, Y. (1995). Controlling the False Discovery Rate: A Practical and Powerful Approach to Multiple Testing. Journal of the Royal Statistical Society. Series B (Methodological), 57(1), 289–300. http://www.jstor.org/stable/2346101

Cohen, M., & Pettit, K. L. S. (2019). Guide to measuring neighborhood change to understand and prevent displacement.

D’Souza, G., Bhondoekhan, F., Benning, L., Margolick, J. B., Adedimeji, A. A., Adimora, A. A., Alcaide, M. L., Cohen, M. H., Detels, R., Friedman, M. R., Holman, S., Konkle-Parker, D. J., Merenstein, D., Ofotokun, I., Palella, F., Altekruse, S., Brown, T. T., & Tien, P. C. (2021). Characteristics of the MACS/WIHS Combined Cohort Study: Opportunities for Research on Aging With HIV in the Longest US Observational Study of HIV. American Journal of Epidemiology, 190(8), 1457–1475. 10.1093/aje/kwab050

Friedman, M. R., Coulter, R. W. S., Silvestre, A. J., Stall, R., Teplin, L., Shoptaw, S., Surkan, P. J., & Plankey, M. W. (2017). Someone to count on: social support as an effect modifier of viral load suppression in a prospective cohort study. AIDS Care, 29(4), 469–480. 10.1080/09540121.2016.1211614

Goldman, A. W., York Cornwell, E., & Cornwell, B. (2023). Neighborhood conditions and social network turnover among older adults. Social Networks, 73, 114–129. 10.1016/j.socnet.2023.01.003

Greene, M., Hessol, N. A., Perissinotto, C., Zepf, R., Hutton Parrott, A., Foreman, C., Whirry, R., Gandhi, M., & John, M. (2018). Loneliness in Older Adults Living with HIV. AIDS and Behavior, 22(5), 1475–1484. 10.1007/s10461-017-1985-1

Hill, T. D., Ross, C. E., & Angel, R. J. (2005). Neighborhood Disorder, Psychophysiological Distress, and Health. Journal of Health and Social Behavior, 46(June), 170–186.

Hughes, M. E., Waite, L. J., Hawkley, L. C., & Cacioppo, J. T. (2004). A Short Scale for Measuring Loneliness in Large Surveys: Results From Two Population-Based Studies. Research on Aging, 26(6), 655–672. 10.1177/0164027504268574

Kamalyan, L., Yang, J.-A., Pope, C., Paolillo, E., Campbell, L., Tang, B., Depp, C., & Moore, R. (2022). SOCIAL INTERACTIONS REDUCE ASSOCIATION BETWEEN LIFE-SPACE AND DAILY HAPPINESS IN OLDER ADULTS WITH AND WITHOUT HIV. Innovation in Aging, 6(Supplement_1), 123. 10.1093/geroni/igac059.492

Lam, J., & Baxter, J. (2025). Neighborhood Characteristics and Loneliness in Later Life: The Role of “Person–Environment Fit.” Innovation in Aging, 9(3), igaf006. 10.1093/geroni/igaf006

Lasgaard, M., Løvschall, C., Qualter, P., Laustsen, L. M., Lim, M. H., Maindal, H. T., Hargaard, A. S., & Christensen, J. (2022). Are loneliness interventions effective in reducing loneliness? A meta-analytic review of 128 studies. *European Journal of Public Health*, *32*(Supplement_3), ckac129.266. 10.1093/eurpub/ckac129.266

Lewinsohn, P. M., Seeley, J. R., Roberts, R. E., & Allen, N. B. (1997). Center for Epidemiologic Studies Depression Scale (CES-D) as a screening instrument for depression among community-residing older adults. Psychology and Aging, 12(2), 277–287. 10.1037/0882-7974.12.2.277

Lucero, C., Sugg, M. M., Ryan, S. C., Runkle, J. D., & Thompson, M. P. (2024). Spatiotemporal patterns of youth isolation and loneliness in the US: a geospatial analysis of Crisis Text Line data (2016-2022). GeoJournal, 89(6), 249. 10.1007/s10708-024-11253-w

Lyu, Yingying, & Forsyth, Ann. (2021). Planning, Aging, and Loneliness: Reviewing Evidence About Built Environment Effects. Journal of Planning Literature, 37(1), 28–48. 10.1177/08854122211035131

Mujahid, M. S., Diez Roux, A. V, Morenoff, J. D., & Raghunathan, T. (2007). Assessing the measurement properties of neighborhood scales: from psychometrics to ecometrics. Am J Epidemiol, 165(8), 858–867. 10.1093/aje/kwm040

Park, S., Smith, J., Dunkle, R. E., Ingersoll-Dayton, B., & Antonucci, T. C. (2019). Health and Social–Physical Environment Profiles Among Older Adults Living Alone: Associations With Depressive Symptoms. The Journals of Gerontology: Series B, 74(4), 675–684. 10.1093/geronb/gbx003

Pearson, A. L., Sadler, R. C., & Kruger, D. J. (2019). Social Integration may Moderate the Relationship between Neighborhood Vacancy and Mental Health Outcomes: Initial Evidence from Flint, Michigan. Applied Research in Quality of Life, 14(4), 1129–1144. 10.1007/s11482-018-9646-8

Pollak, C., Cotton, K., Winter, J., & Blumen, H. (2025). Health Outcomes Associated with Loneliness and Social Isolation in Older Adults Living with HIV: A Systematic Review. AIDS and Behavior, 29(1), 166–186. 10.1007/s10461-024-04471-3

Radloff, L. L. S. (1977). The CES-D scale a self-report depression scale for research in the general population. Applied Psychological Measurement, 1(3), 385–401. 10.1177/014662167700100306

Raudenbush, S. W., & Sampson, R. J. (1999). Ecometrics: Toward a Science of Assessing Ecological Settings, with Application to the Systematic Social Observation of Neighborhoods. Sociological Methodology, 29, 1–41.

Ross, C. E. (2000). Neighborhood Disadvantage and Adult Depression. Journal of Health and Social Behavior, 41(June), 177–187.

Ross, C. E., & Mirowsky, J. (2001). Neighborhood Disadvantage, Disorder, and Health. Journal of Health and Social Behavior, 42(September), 258–276.

Ross, C. E., & Mirowsky, J. (2009). Neighborhood Disorder, Subjective Alienation, and Distress. Journal of Health and Social Behavior, 50(March), 49–64.

Ross, C. E., Reynolds, J. R., & Geis, K. J. (2000). The Contingent Meaning of Neighborhood Stability for Residents’ Psychological Well-Being. American Sociological Review, 65(4), 581–597. 10.2307/2657384

Russell, D., Peplau, L. A., & Ferguson, M. L. (1978). Developing a Measure of Loneliness. Journal of Personality Assessment, 42(3), 290–294. 10.1207/s15327752jpa4203_11

Scheidt, R. J., Norris-Baker, C., Wahl, H. W., & others. (2003). The general ecological model revisited: Evolution, current status, and continuing challenges. Annual Review of Gerontology and Geriatrics, 23, 34–58.

Schieman, S. (2009). RESIDENTIAL STABILITY, NEIGHBORHOOD RACIAL COMPOSITION, AND THE SUBJECTIVE ASSESSMENT OF NEIGHBORHOOD PROBLEMS AMONG OLDER ADULTS. The Sociological Quarterly, 50(4), 608–632. 10.1111/j.1533-8525.2009.01158.x

Singer, J. D., & Willett, J. B. (2003). Applied longitudinal data analysis: Modeling change and event occurrence. Oxford university press.

Wahl, H.-W., & Lang, F. R. (2003). Aging in context across the adult life course: Integrating physical and social environmental research perspectives. Annual Review of Gerontology and Geriatrics, 23, 1–33.

Wu, A. W., Hays, R. D., Kelly, S., Malitz, F., & Bozzette, S. A. (1997). Applications of the Medical Outcomes Study health-related quality of life measures in HIV/AIDS. Quality of Life Research, 6(6), 531–554. 10.1023/A:1018460132567

Zhao, T., Tang, C., Ma, J., Halili, X., Yan, H., & Wang, H. (2025). Interventions for subjective and objective social isolation among people living with HIV: A scoping review. Social Science & Medicine, 367, 117604. 10.1016/j.socscimed.2024.117604

